# Publicly available continuously updated topic specific databases of randomised clinical trials: A scoping review

**DOI:** 10.1101/2024.11.18.24317477

**Authors:** Kim Boesen, Lars G Hemkens, Perrine Janiaud, Julian Hirt

## Abstract

Conducting systematic reviews of clinical trials is arduous and resource consuming. One potential solution is to design databases that are continuously and automatically populated with clinical trial data from harmonised and structured datasets. We aimed to map publicly available, continuously updated, topic-specific databases of randomised clinical trials (RCTs). We systematically searched PubMed, Embase, the preprint servers medRxiv, ArXiv, and Open Science Framework, and Google. We described seven features (access model, database architecture, data input sources, retrieval methods, data extraction methods, trial presentation, and export options) and narratively summarised the results. We did not register a protocol for this review. We identified 14 continuously updated clinical trial databases, seven related to COVID-19 (first active in 2020) and seven non-COVID databases (first active in 2009). All databases, except one, were publicly funded and accessible without restrictions. They mainly employed methods similar to those from static article-based systematic reviews and retrieved data from journal publications and trial registries. The COVID-19 databases and some non-COVID databases implemented semi-automated features of data import, which combined automated and manual data curation, whereas the non-COVID databases mainly relied on manual workflows. Most reported information was metadata, such as author names, years of publication, and link to publication or trial registry. Two databases included trial appraisal information (risk of bias assessments). Six databases reported aggregate group level results, but only one database provided individual participant data on request. We identified few continuously updated trial databases, and existing initiatives mainly employ methods known from static article -based reviews. The main limitation to create truly live evidence synthesis is the access and import of machine-readable and harmonised clinical trial data.

## Introduction

The number of conducted and published clinical trials and systematic reviews has increased tremendously over the past decades.^1, 2^ It is difficult, if not impossible, for any clinician or researcher to stay up to date. Traditional systematic reviews are time consuming and costly to produce due to the time required to manually search databases, screen titles, assess studies, and extract data.^3^ Systematic reviews are often outdated and potentially even misleading when published. ^4-7^ Different tools have been launched to automatise the review conduct and maintenance to reduce the costs and resources.^8-11^ Those tools focus mainly on text mining and machine learning assistance to screen and retrieve records^12^ and extract data from published reports of clinical trials.^13^

Due to the systematic reviews’ limitations of staying up to date, the concept of ‘living systematic reviews’ has popularised. There is no clear definition of what a ‘living review’ is other than it is a (systematic) review being continuously updated.^14,15^ In theory it should mitigate delays and the risk of missing important evidence. In 2017, the Cochrane Collaboration published what they call “the world’s first ‘living systematic review”. The review was published in 2017, and it has still not being updated in the Cochrane Library.^16,17^ In recent years, the COVID-19 pandemic ignited a torrent of such ‘live’ projects reflecting the rapidly evolving evidence, including the COVID-19 trial tracker,^18^ the NMA-Covid Project,^19^ and WHO’s living guideline^20^ that were known to us prior to this scoping review (S1, Appendix).

The first requirement to enable such continuously updated databases of clinical trial data is to have all trial data available in a machine-readable format and structured in a harmonised fashion.^21^ The idea of such machine-readable repository of RCTs goes 25 years back with the Global Trial Bank. It was a system envisioned to go beyond trial registration to also contain trial results reporting in a computable fashion.^22-25^ The Global Trial Bank never materialised, but along these lines the International Committee for Journal Medical Editors’ clinical trial registry requirement as a pre-empt for publication came into place in 2004.^26^ Subsequently, it also became mandatory to submit summary results on ClinicalTrials.gov (2007) and on the European Union Clinical Trial Register (2012).^27^

Some researchers have voiced that clinical trial data should be made available in a structured dataset, like the Common Technical Document format used for commercial drug applications.^28^ Others have argued that evidence synthesis, like systematic reviews, should rely primarily on clinical trial registries as the main data source, rather than the traditional decentralised publication-based system, to enable more timely and exhaustive evidence synthesis.^29^ The concept of the Global Trial Bank and of continuously updated, clinical trial databases lies in the intersection of these two ideas.

Whether existing living projects and topic specific trial databases leverage advanced technologies to ensure seamless and – ideally fully automated – flow of clinical trial data into structured databases is unknown. We therefore aimed to systematically identify publicly available continuously updated topic-specific databases of clinical trials to characterise basic functionalities, infrastructure, and user interface. We used this work as a primer for the design of a continuously updated database on specific cancer immunotherapies.^30^

## Methods

We conducted a scoping review and due to its exploratory nature, we did not follow a protocol or register this review since we anticipated numerous unforeseeable challenges during mapping this field. We report this scoping review according to the Preferred Reporting Items for Systematic reviews and Meta-Analyses statement for Scoping Reviews (PRISMA-ScR).^31^

### Eligibility criteria

We included any publicly available continuously updated database retrieving RCTs (participants allocated to one of two or more groups); irrespective of retrieval method (e.g. manually, automated, or a combination), input source (e.g. databases of published literature, trial registries, or both), and we did not define a minimum update frequency threshold. We considered field or topic specific databases only, i.e. exclusive to one condition, specialty, intervention, or indication, such as a COVID-19. Databases that also retrieved other research designs, such as observational studies, were still considered eligible. We did not include broad, unspecific trial databases, such as clinical trial registries. We applied no restrictions on language, release date, intended use (i.e., for research or clinical decision-making), or the database status (i.e. live, archived, or unknown status).

We did not include conventional, static, article-based ‘living systematic reviews’ without an eligible database adhering to the inclusion criteria above.

### Information sources and search strategy

We employed a two-step systematic search strategy. First, we did a “cold search” of various resources to identify key words and seed references to subsequently build a systematic database search.

#### Step 1: Cold (‘seed’) search

One author (KB) searched PubMed (including using the “similar articles” function), preprint servers medRxiv and ArXiv, Open Science Framework, and Google (first five pages for each search) in January 2023 (S2, Appendix).

#### Step 2: Systematic search

Two authors (KB, JH) designed a three-component search strategy containing keywords for “living” (i.e. live, up-to-date, updated, continuously updated, accumulating, dynamic, digital, and interactive), keywords for “library” (i.e. database, platform, hub, repository, collection, overview, archive, body of evidence, index, bibliography, catalogue, compilation, portal, inventory, and map), and study design (i.e. RCT, trial, and clinical study). One author (JH) searched PubMed and Embase (S3 and S4, Appendix) on February 1, 2023, deduplicated search results using Citavi,^32^ and imported the results into the screening tool Rayyan.^33^ One author (KB) screened titles, abstracts, and full texts and decided on inclusion in one step.

### Data extraction and analysis

We described and narratively summarised the following seven database features. One reviewer (KB) extracted information, and one reviewer (JH) double-checked and confirmed the extractions.

#### Access model

How is the database content accessed? Unrestricted access (i.e. data is freely available without any constraints), controlled access (e.g. users have to submit requests and the provider grants or denies access), or a hybrid between the two.

#### Database architecture

What is the underlying database structure and used database management software? Simple Excel spreadsheet or relational databases,^34^ and were proprietary data management tools used, like RedCap.^35^

#### Input sources

Which data input sources were searched? Databases of published literature like PubMed or Embase, trial registries, e.g. ClinicalTrials.gov, or other sources.

#### Retrieval methods

How is the database populated with trials? Manually, automated e.g. by application programming interfaces (APIs), or community-based data submissions, like ClinicalTrials.gov or the nucleotide sequencing database GenBank, where researchers submit data to the database.^36^

#### Data extraction

How is the trial information extracted and curated? Manually, fully automated, or a combination of machine-assisted manual curation, usually referred to as ‘semi-automated’.

#### Trial presentation

What trial information are provided? Meta data (e.g. title, author names, publication or trial registry information), detailed trial information (e.g. study design, sample size, treatment descriptions, funding), results (outcomes and effect estimates), type of results (aggregate group level or individual participant data), and trial appraisal (e.g. risk of bias assessments, limitations in the trial design).

#### Export options

What are the options for downloading and reusing data? Free download of the dataset (e.g. as a CSV file), federated access ^37^ where users can view and work with the data remotely, e.g. in browser-based applications without downloading it to their own personal computer such as Vivli.com,^38^ or no option for reusing data.

## Results

### Search results

Our systematic database search returned 1707 hits (deduplicated), of which 15 were assessed in full-text from which one database (TrialsResultsCenter^39-40^) was included. One database (Evidence Finder^41-43^) was found through other sources, nine databases (Cochrane COVID-19 study register,^44^ COVID-evidence,^45^ COVID TrialsTracker,^18^ COVID-NMA initiative,^19^ EPPI Centre Covid-19 Living Map of the Evidence,^46^ MetaEvidence breast cancer,^47^ MetaEvidence COVID,^48^ MetalO,^49^ and MetaPreg^50^) were known to us before our systematic search, and our cold searches yielded three other eligible databases (Infectious Disease Data Observatory (IDDO),^51^ Worldwide Antimalarial Resistance Network (WWARN),^52-54^ and EvidenceMap^55-57^; S5, Appendix).

In total, we included 14 databases (Table 1), seven non-COVID databases on youth mental health (Evidence Finder), pancreatic surgery (Evidence Map), cardiology and oncology trials (TrialResultsCenter), cancer (MetaEvidence breast cancer and MetalO), pregnancy (MetaPreg), and malaria (WWARN), and seven COVID-19 databases (Cochrane COVID-19 study register, COVID-evidence, COVID TrialsTracker, COVID-NMA initiative, EPPI Centre Covid-19 Living Map of the Evidence, IDDO, and MetaEvidence COVID).

**Table 1.** Included continuously updated field-specific clinical trials databases.

| Name | Year | Field | Funding/responsible organisation | Access |
| --- | --- | --- | --- | --- |
| **Cochrane COVID-19 study register** | 2020-2024 | COVID | Cochrane and the Federal Ministry of Education and Research of Germany (CEOsyst project) | Fully open |
| **COVID-evidence** | 2020-2022 | COVID | Swiss National Science Foundation | Fully open |
| **COVID NMA initiative** | 2020-2023 | COVID | Centre for Research in Epidemiology and Statistics (CRESS) and Cochrane France | Fully open |
| **COVID Trials Tracker** | 2020-2022 | COVID | Centre for Evidence Based Medicine, Oxford University | Fully open |
| **EPPI Centre Covid-19 Living Map of the Evidence** | 2019-2023 | COVID | Evidence for Policy and Practice (EPPI) Centre and the UK National Institute for Health and Care Research (NIHR) Policy Research Programme Reviews Facility | Fully open |
| **Evidence Finder** | 2010 - 2021 | Youth mental health | Orygen Ltd<sup>a</sup> | Fully open |
| **Evidence Map** | 2021 – active | Pancreatic surgery | International Study Group for Pancreatic Surgery and Viatrix<sup>b</sup> | Fully open |
| **IDDO Infectious disease Data Observatory Living systematic review** | 2020 - active | COVID | Coalition for Equitable Research in Low-resource Settings (CERCLE)<sup>c</sup> | Fully open |
| **MetaEvidence breast cancer** | Not stated | Breast cancer | EU Horizon 2020 Qualitop, Hospices Civils de Lyon, CNRS UMR5558 LBBE | Fully open |
| **MetaEvidence COVID** | 2020-2022 | COVID | Hospices Civils de Lyon | Fully open |
| **MetaIO** | Not stated | Cancer | EU Horizon 2020 Qualitop, Hospices Civils de Lyon, CNRS UMR5558 LBBE | Fully open |
| **MetaPreg** | Not stated | Pregnancy | ANSM (agence Nationale de Sécurité du Médicament), Hospices Civils de Lyon and University Lyon-1 (LBBE) | Fully open |
| **TrialResultsCenter** | 2009 - 2017 | Cardiology and oncology | Hospices Civils de Lyon | Fully open |
| **WWARN Worldwide Antimalarial Resistance Network Clinical Trials Publication Library** | 2011 - active | Malaria | Coalition for Equitable Research in Low-resource Settings (CERCLE)<sup>d</sup> | Fully open |

a) An Australian charity devoted to mental health issues (https://www.orygen.org.au/).
b) We could not find information on the International Study Group for Pancreatic Surgery including its funding. Viatrix is a pharmaceutical company.
c) An international research network currently funded by the German Ministry of Education and Research.
d) WWARN is one of IDDO's projects, thus we assume the funding is the same.

### Basic database characteristics (non-COVID)

TrialResultsCenter was active from 2009 until 2017 (and is no longer updated), EvidenceFinder was released in 2010 (last update in July 2021, uncertain if it is still maintained), WWARN was released in 2011 (uncertain if it still maintained; the last trial in their Data Inventory^52^ dates from 2018 but their trial summary^51^ says it is current until 2022), and EvidenceMap was released in 2021 (last update 26 July 2023). For three databases (MetaEvidence breast cancer, MetalO, and MetaPreg) the start was not stated. Six of the non-COVID databases were either publicly funded or funded by a non-profit organisation, and one (Evidence Map) was funded by a pharmaceutical company. All database websites were fully accessible with no restrictions.

The databases generally searched common sources, mainly databases of published reports including PubMed/MEDLINE, Embase, and Web of Science, using methods resembling those of regular systematic reviews with manual database searches, extraction, and data curation. EvidenceMap was built on a relational Microsoft SQL server database (Windows Form .NET management system), WWARN uses a proprietary data management tool (RedCap), and it was not described for the other five. Five databases (EvidenceMap, MetaEvidence breast cancer, MetalO, MetaPreg, and WWARN) made their data readily available for download, one (Trial Results Center) upon request, and one (Evidence Finder) did not mention options for download (Table 2).

**Table 2.** Database basic characteristics (non-COVID)

| Name | Database architecture | Input sources | Retrieval methods (frequency) | Curation | Trial presentation | Export |
| --- | --- | --- | --- | --- | --- | --- |
| **Evidence Finder** | Not described | MEDLINE, Embase, PsychINFO, Cochrane Library | Manually (yearly) | Manual | Metadata, trial information | Not mentioned |
| **Evidence Map** | Relational database (Windows Form .NET management system) | PubMed, CENTRAL, Web of Science | Manually (frequency not described) | Manual | Metadata, trial appraisal | Excel download |
| **MetaEvidence Breast cancer** | Not described * | Not described * | Not described * | Not described * | Metadata, trial information, trial appraisal, results (group level) | No option |
| **MetalO** | Not described * | Not described* | Not described* | Not described* | Metadata, trial information, trial appraisal, results (group level) | No option |
| **MetaPreg** | Not described ("Proprietary meta-analysis platform") | PubMed, Scopus, Web of Science, 17 preprint servers | Semi-automated (daily) ("using automatic software robots") | Manual | Metadata, trial information, trial appraisal, results (group level) | No option |
| **Trial Results Center** | Not described | MEDLINE, Embase, CENTRAL, ClinicalTrials.gov, ISRCTN, ICTRP | Manually (weekly) | Manual | Metadata, trial information, results (group level) | On request |
| **WWARN** | Proprietary data management system (RedCap) | PubMed, Embase, Web of Science | Manually (every 6 months) | Manual | Metadata | Excel download |

There is no separate method s description for these platforms. They link to the metaEvidence COVID methods.

All databases provide basic metadata information, e.g. title, author names, and link to publication/trial registry. Three databases (MetaEvidence breast cancer, MetalO, and MetaPreg) reported aggregated group level results and risk of bias assessments, one (TrialsResultsCenter) reported aggregated group level results, two (Evidence Finder and TrialsResultsCenter) reported some trial information, and one (EvidenceMap) reported trial appraisal in the shape of risk of bias assessment (Table 2; S6, Appendix).

### Basic database characteristics (COVID)

The seven COVID databases were initiated shortly after the SARS-COV-2 outbreak in early 2020, and by January 2024 the last database, the Cochrane COVID-19 study register, stopped being updated. They were all publicly funded or funded by non-profit institutions and open to the public. All websites were fully accessible with no restrictions.

Three of the seven COVID databases (Cochrane COVID-19 study register, EPPI Centre Covid-19 Living Map of the Evidence, and MetaEvidence COVID) generally searched common sources, mainly databases of published reports including PubMed/MEDLINE and Embase. Six of the seven databases searched (in addition) one or more trial registries. Three databases (Cochrane COVID-19 study register, COVID-evidence, COVID TrialsTracker, EPPI Centre Covid-19 Living Map of the Evidence, and Meta-evidence) used semi-automated methods for retrieval of studies and data extraction, mainly for trial registry records, and two (COVID NMA Initiative and IDDO) manually retrieved and extracted data only. Two (IDDO and COVID-evidence) used a proprietary management tool (RedCap and Directus), and for the others it was not described. Six databases (Cochrane COVID-19 study register, COVID-evidence, COVID TrialsTracker, EPPI Centre Covid-19 Living Map of the Evidence, MetaEvidence COVID, and IDDO) made their data available for download and one (COVID NMA initiative) made it available upon request (Table 3).

**Table 3.**
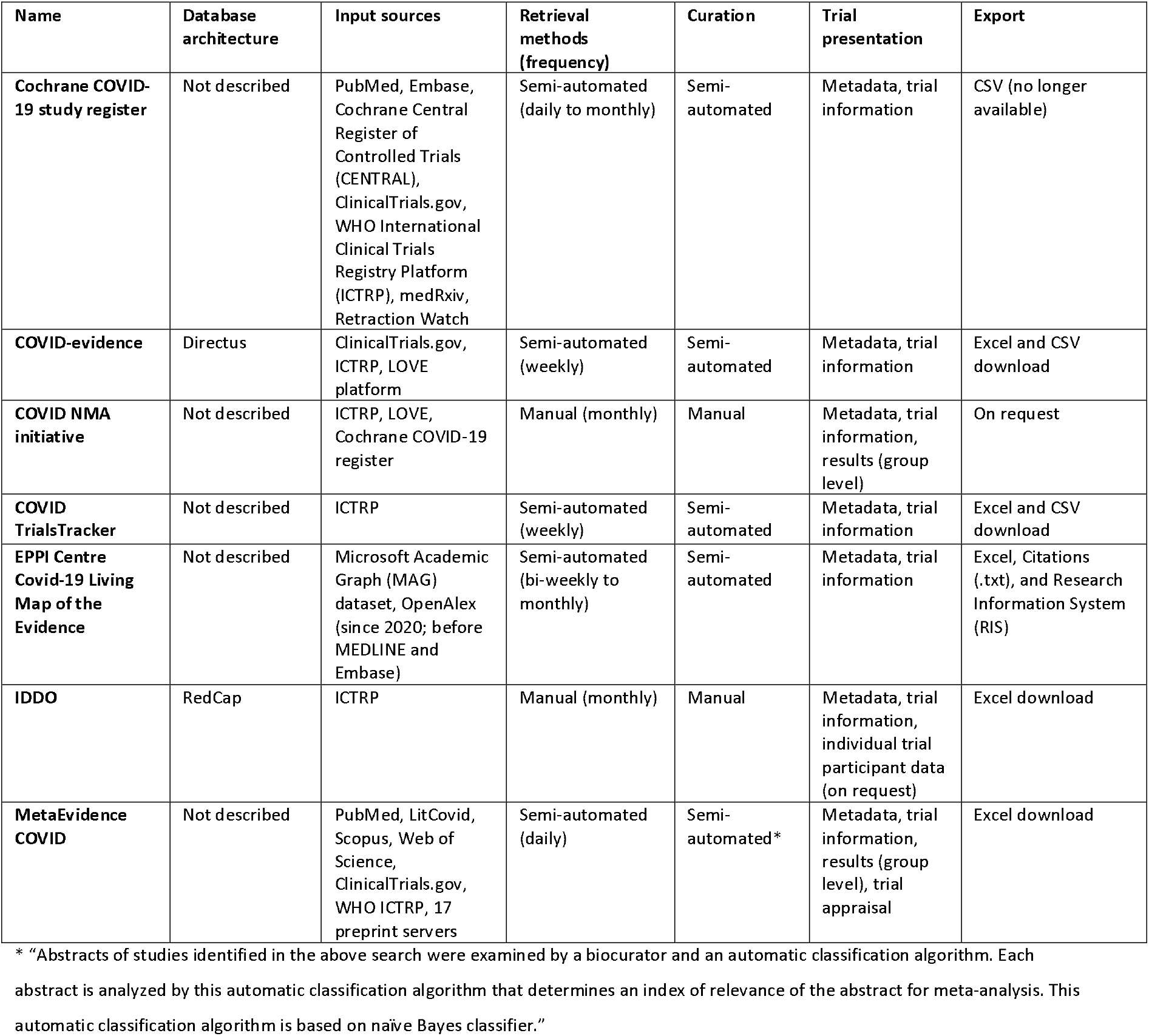
Database basic characteristics (COVID specific)

All databases reported basic metadata and also trial information (such as study type or design, sample size or expected starting date extracted from ClinicalTrials.gov), two (Covid NMA Initiative and MetaEvidence COVID) reported aggregate group level results, and one (MetaEvidence COVID) also reported trial appraisal in the shape of risk of bias assessments (Table 3; S6, Appendix).

## Discussion

We identified 14 continuously updated clinical trials databases, of which 7 were COVID-19 related. The COVID projects used semi-automated methods to retrieve and extract data, primarily from clinical trial registries, whereas the non-COVID databases mainly employed manual methods from regular systematic reviews, which are time consuming and difficult to maintain. All databases presented mainly metadata whereas curated trial information, results, or trial appraisal were sparsely reported. Only the IDDO database explicitly stated to provide the option of sharing individual participant data, which can then be accessed on request.

### Findings in context

Our scoping review highlights a dearth of continuously updated topic-specific trial databases. Existing databases borrow methods and technology from static article-based systematic reviews. The main issue seems that most information from published clinical trials must be retrieved, extracted, and curated from unstructured journal articles, and that data captured or exported from clinical trial registries are not easily incorporated into databases. None of the 14 platforms solved this problem, for instance by retrieving data from other sources such as standardised machine-readable datasets. We do not have knowledge of such publicly available structured datasets. The public-private FDA Critical Path Initiative ^58^ has established databases of Alzheimer’s,^59,60^ multiple sclerosis ^61^, Parkinson’s ^62^ and tuberculosis ^63^ clinical trial data, although these are not publicly available. The prevalence of such public-private or fully private data platforms is unknown to us.

There are several reasons why existing trial registries are challenging as a data source for creating topic-specific trial databases. First of all, the various trial registries do not harmonise database schemes or data format structures beyond the WHO Trial Registration Data Set.^64^ It is a high-level reporting checklist that do not specify exhaustive outcome specifications and results reporting. Secondly, based on our initial experiences with the CIEL database,^30^ we assume that the error rates of inserting wrong data or into the wrong fields in clinical trial registries is very high. This makes the usefulness of machine-readable trial registry datasets (e.g. ClinicalTrials.gov’s dataset that is accessible through an application programming interface or API), rather limited. Imported data needs a substantial amount of “data cleaning” to become useful. Thirdly, only a few registries enable data export, whereas others, such as the European Union Clinical Trial Register does not have an option for automated access to data.

One potential tool to solve the problem of extracting data from journal publications is TrialStreamer.^65^ It is not a topic-specific database, but a mega-collection of PubMed indexed clinical trials (n = 852.723 by 20 Dec 2023), with annotated PICO information (population, information, comparator, outcome). Trial Streamer uses a mix of machine-learning and crowd-sourced manual extraction to retrieve the data. TrialStreamer may be useful for high-level meta-epidemiological studies, but for clinical applications or guideline development, more granular trial descriptions are needed.

The importance of living evidence synthesis was recently acknowledged by the Wellcome Trust.^66^ They will invest GBP 45 million over a five-year period to establish and accelerate the development of data infrastructure for future living evidence projects.

### Limitations

Our scoping review has limitations. First, we did not prespecify our methodology and register a protocol for this work but we believe that the impact is minimal considering the scope of the review with itsexploratory nature. Second, we have likely missed datasources due to the scarcity and disparity of databases, lacking common terminology, and lacking related published articles. We are confident that many ‘stealth’^67^ commercial solutions exist. An example of a public commercial solution is the LiveSLR program,^68,69^ which is a software combining an annotated library of publications (called LiveRef) with a machine-assisted search/screening/ranking workflow, which creates an interactive live platform used in drug applications and health technology dossiers. Unfortunately, there is little transparency when it comes to such commercial solutions, including LiveSLR, and we cannot describe such solutions in more details. Finally, we decided to not make detailed assessments of the database infrastructure, such as granular descriptions of how data was retrieved and extracted automatically from ClinicalTrials.gov. Mainly because it was not transparently reported on the websites (nor appeared from the journal publications that we found in our systematic search). Transparent and exhaustive reporting of the database infrastructure should be a priority because automation is the main methodological challenge. For example, the metaPreg platform described to use “automatic software robots” to screen databases, with no further specifications.^50^ We acknowledge that there may be several reasons for vaguely describing the methodology, for example because such setups are likely updated and adjusted frequently. However, open and complete descriptions of platform infrastructure, including how automation is attempted and achieved, is the fastest way to advance this field.

## Conclusions

This is the first review on the topic of continuously updated clinical trial databases. It could inform the design of future databases and outline technical and methodological challenges. We found only few topic-specific databases of clinical trials, which mainly relied on manual searches and manual data extraction. Such approaches are time consuming and inefficient to keep the databases updated. Recent COVID-19 trial databases implemented semi-automated workflows, albeit confined to clinical trial registries. A major challenge remains how to automatically extract trial data from journal publications. It seems timely to reconsider ideas such as the Global Trial Bank to centralise, not only trial registration, but also trial results reporting and data sharing. Facilitating the shift from article-based publications to a fully ‘digital’ machine-readable system of clinical trial data could enable true live evidence synthesis.

## Data Availability

All data related to this review are available in the paper and the Appendix.

## Acknowledgements

None.

## Authors’ contributions

KB, PJ, and JH made substantial contributions to the conception and design of the work; KB have drafted the work and all authors substantively revised it; all authors have approved the submitted version; all authors have agreed both to be personally accountable for the author’s own contributions and to ensure that questions related to the accuracy or integrity of any part of the work, even ones in which the author was not personally involved, are appropriately investigated, resolved, and the resolution documented in the literature.

## Data sharing statement

All data related to this review are available in the paper and the Appendix.

