## Appendix for "Publicly available continuously updated topic specific databases of randomised clinical trials: A scoping review"

Boesen et al.

### Appendix

S1. Clinical trial databases known before scoping review

S2. Cold searches to identify seed references and key words

S3. Search string (PubMed)

S4. Search string (Embase)

S5. PRISMA 2020 flowchart of the literature search and selection process

S6. Screenshots of trial presentations

### S1. Clinical trial databases known before the scoping review

| Platform | Funder | Type of platform | Comments |
|---|---|---|---|
| Cochrane COVID-19 study register https://covid-19.cochrane.org/ | Cochrane Collaboration | Database of COVID studies, including but not restricted to RCTs | - |
| COVID-evidence https://covid-evidence.org/ | Swiss National Science Foundation | Database of COVID RCTs | Not updated since 31/12/2022 |
| COVID-NMA initiative https://covid-nma.com/ | CRESS and Cochrane France | Live mapping and network meta-analyses of COVID trials | - |
| COVID 19 Trial Tracker https://covid19.trialstracker.net/ | CEBM, Oxford University | Database of COVID RCTs | - |
| EPPI Centre Covid-19 Living Map of the Evidence https://eppi.ioe.ac.uk/eppi-vis/Review/Index/7 | EPPI Centre | Database of COVID studies, including but not restricted to RCTs | - |
| MetaEvidence COVID http://www.metaevidence.org/default0.aspx | Hospices Civils de Lyon | Database of COVID RCTs | No longer updated |
| MetaEvidence breast cancer http://www.metaevidence.org/default0.aspx | Hospices Civils de Lyon | Database of breast cancer treatments | - |
| MetaIO http://www.metaevidence.org/default0.aspx | Hospices Civils de Lyon | Database of immuno-oncology trials | - |
| PragMeta* https://pragmeta.org/ | Swiss National Science Foundation | Database of pragmatic trials | Not continuously updated |

*Not qualifying as a 'continuously updated field-specific databases'; either not continuously updated or not clinical trial or field specific, or unable to assess the used methods (no descriptions, or offline).

S2. Cold searches to identify seed references and key words

| Source | Search string | No. hits | Potential relevant hits |
|---|---|---|---|
| **Pubmed** | “living library” | 8 | 0 |
| | “living review”[a] | 67 | 0 |
| | “living systematic review”[b] | 276 | 1 (Evidence map) |
| | “Living evidence map” | 4 | 1 (Evidence map) |
| | “Living platform” | 6 | 0 |
| | “Living database” | 11 | 1 (Trial streamer) |
| | “Clinical trials platform” | 82 | 0 |
| | “interactive database”[c] | 258 | 0 |
| **PROSPERO** | “Living library” | 1 | 0 |
| | “Interactive database” | 0 | 0 |
| | “Clinical trials platform” | 80 | 0 |
| **medRxiv** | “Living library” | 0 | 0 |
| | “Living review” | 84 | 0 |
| | “Evidence map” | 49 | 1 (Trialstreamer) |
| | “Database clinical trials” | 12 | 0 |
| **ArXiv** | “information retrieval” AND “medical” AND ”trial” (field codes) | 9 | 1 (TrialStreamer) |
| **OSF** | “Living library” | 0 | |
| | “Living database” (project) | 6 | 1 (Covid evidence) |
| | “Evidence map” (project) | 37 | 0 |
| | “Clinical trials platform” | 2 | 0 |
| **Google** | “Living library clinical trials”, “library clinical trials”, “living platform clinical trials” | Infinitive | 3 (Atlas Edu, IDDO, WWARN) |
| **Other sources** | - | - | 1 (Evidence Finder) |

a) Mostly regular “paper based” living systematic reviews.
b) Mostly regular systematic reviews, majority about COVID.
c) Many hits related to genomic databases, such as https://doi.org/10.1093/nar/gkz884 (database for snoDNA); https://doi.org/10.3389/fgene.2019.00262 (database for human single nucleotide polymorphism SNP).

S3. Search string (PubMed)

### MEDLINE/PubMed

Search Feb 1, 2023:

| # | Entry | Hits |
|---|---|---|
| 1 | (database*[title] OR librar*[title] OR platform*[title] OR hub[title] OR repositor*[title] OR collection*[title] OR overview*[title] OR archive*[title] OR index*[title] OR bibliograph*[title] OR catalogue*[title] OR system[title] OR storage*[title] OR data*[title] OR retrieval*[title] OR inventor*[title] OR portal[title]) AND (living[title] OR up-to-date[title] OR updat*[title] OR continuous*[title] OR accumulat*[title] OR dynamic[title] OR interactive[title] OR digital[title]) AND (rct[tiab] OR trial[tiab] OR "clinical study"[tiab] OR "Clinical Studies as Topic"[MeSH]) | 1,079 |

S4. Search string (Embase)

### Embase/Elsevier

Search Feb 1, 2023:

| # | Entry | Hits |
|---|---|---|
| 1 | (database*:ti OR librar*:ti OR platform*:ti OR hub:ti OR repositor*:ti OR collection*:ti OR overview*:ti OR archive*:ti OR index*:ti OR bibliograph*:ti OR catalogue*:ti OR system:ti OR storage*:ti OR data*:ti OR retrieval*:ti OR inventor*:ti OR portal:ti) AND (living:ti OR 'up to date':ti OR updat*:ti OR continuous*:ti OR accumulat*:ti OR dynamic:ti OR interactive:ti OR digital:ti) AND (rct:ti,ab OR trial:ti,ab OR 'clinical study':ti,ab OR 'clinical study'/de) | 1,507 |
| 2 | #1 AND [embase]/lim NOT ([embase]/lim AND [medline]/lim) | 725 |

S5. PRISMA 2020 flowchart of the literature search and selection process

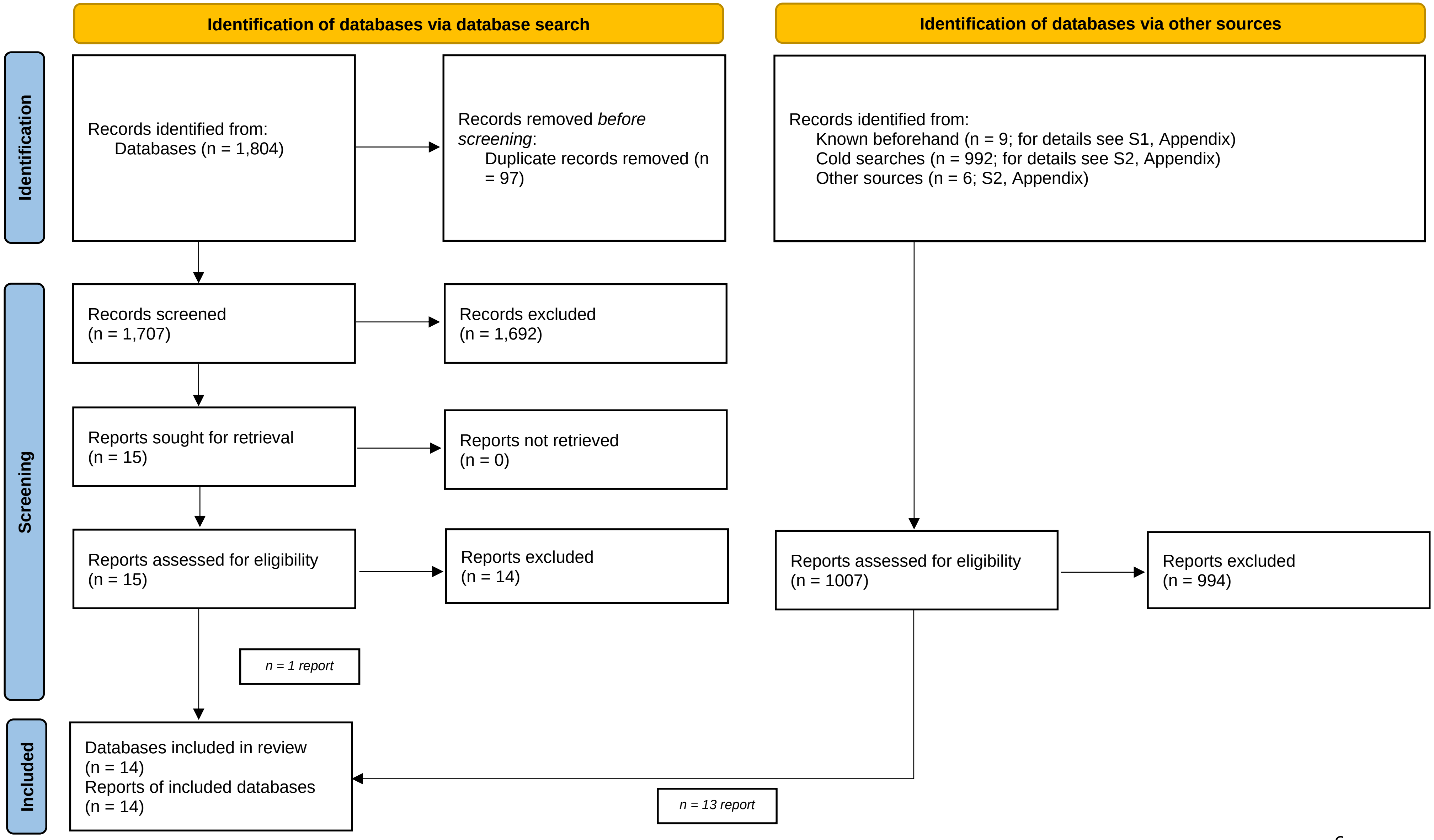

### S6. Screenshots of trial presentations

#### Cochrane COVID-19 study register

STUDY RECORD

MURAKAMI, 21 OCT 2023 | First Published: 2024 Feb 4 | Updated Date: 2024 Feb 4

**Perceptions and responses to COVID-19 through wastewater surveillance information and online search behavior: A randomized controlled trial**

| Study Type | Study Aim | Study Design | Intervention Assignment |
|---|---|---|---|
| Interventional | Epidemiology | Parallel/Crossover | Randomised |

REFERENCE RECORD

**Perceptions and responses to COVID-19 through wastewater surveillance information and online search behavior: A randomized controlled trial**

Murakami, M.; Nomura, S.; Ando, H.; Kitajima,,M...
First Published | ↗ Medrxiv (https://www.medrxiv.org/content/early/2023/10/21/2023.10.20.23297297)
Unpublished article (preprint)
Report Results

Wastewater surveillance is anticipated to be a representative and timely method to assess infectious disease status; however, its influence on public perception and behavior remains unclear. Therefore, in this study, we used a randomized controlled trial to analyze the influence of wastewater surveillance-based information on understanding of, interest in, relief regarding, preventive behavioral intention against, and subsequent online search behavior related to coronavirus disease 2019 (COVID-19). Valid responses were obtained from 1,000 individuals in both control and intervention groups from Yahoo crowdsourcing users aged [≥]18 years in Japan. This survey was conducted from August 4 to August 7, 2023, just before the common Japanese tradition of returning to hometowns. The questionnaire not only collected personal attributes but also gauged responses to COVID-19 information. This information highlighted the early detection capabilities and representativeness of wastewater surveillance compared with sentinel surveillance at medical institutions. At one-week post-survey, we obtained the survey participants' online search history for key words such as "bullet train," "highway," "airplane," and "wastewater." The findings showed no significant differences between the two groups in terms of COVID-19 interest or preventive behavior before information provision, verifying the effectiveness of participant randomization. Wastewater surveillance-based information did not notably elevate understanding or specific intentions regarding COVID-19, such as wearing masks and receiving vaccination. However, it significantly increased interest in, relief concerning the infection status, and general preventive behavioral intentions. Heightened interest and general preventive intentions did not depend on prior interest or behavior. However, those who previously engaged in preventive behavior or who were less interested in COVID-19 exhibited more relief after exposure to wastewater surveillance-based information. Furthermore, this information could slightly influence online searches related to return travel modes, such as highways. In conclusion, information from wastewater surveillance effectively shapes individual perceptions of and responses to infections. [Preprints are preliminary reports of work that have not been peer reviewed. Refer to the original preprint or preprint server for specific information about the individual preprint.]

#### COVID Evidence

- **Bringing Optimised COVID-19 Vaccine Schedules To ImmunoCompromised Populations (BOOST-IC): an Adaptive Randomised Controlled Clinical Trial**

  *NCT05556720 Location: Australia (Australia/Oceania) Status: Not Yet Recruiting Registration Date: 2022-09-27 Start Date: 2022-10-01 Estimated End Date: 2025-12-31*

  Brief summary: Despite the greater risk of adverse COVID-19 outcomes, antibody and cell-mediated immune responses to COVID-19 vaccines vary amongst immunocompromised (IC) people and are poorly defined. IC hosts were largely excluded from the COVID-19 vaccine registration trials, though many countries...

  @clinicaltrials.gov

#### COVID NMA initiative

| Treatment (per arm) | Sample size | Severity at enrollment | Sponsor/Funder | Reg. number |
|---|---|---|---|---|
| (1) Nirmatrelvir + ritonavir *vs* (2) Nirmatrelvir + ritonavir *vs* (3) Placebo | 900 | Patients recovered from covid | Kanecia Obie Zimmerman | NCT05595369 |
| (1) Imidazolyl ethanamide pentandioic acid *vs* (2) Placebo | 233 | Mild | Valenta Pharm JSC | NCT05216497 |
| (1) Isotretinoin *vs* (2) Isotretinoin *vs* (3) Standard of care | 100000 | Critical | Kafrelsheikh University | NCT04353180 |
| (1) Interferon gamma *vs* (2) Standard of care | 630 | Healthy volunteers | SPP Pharmaclon Ltd. | NCT05054114 |
| (1) Fluticasone *vs* (2) Standard of care | 500 | Mild | University of Medicine and Pharmacy at Ho Chi Minh City | NCT05054322 |
| (1) Baricitinib *vs* (2) Placebo | 480 | Moderate/severe | Incepta Pharmaceuticals Ltd | NCT05056558 |
| (1) Platelet rich plasma *vs* (2) Placebo | 30 | Patients recovered from covid | Stanford University | NCT04406584 |
| (1) Masitinib *vs* (2) Masitinib *vs* (3) Masitinib *vs* (4) Placebo | 78 | Mild/moderate | AB Science | NCT05047783 |
| (1) Isotretinoin + tamoxifen *vs* (2) Isotretinoin + tamoxifen | 160 | Severe/critical | Kafrelsheikh University | NCT04389580 |
| (1) Placebo | 400 | Healthy volunteers | Chumakov Federal Scientific Center for Research and Development of Immune-and-Biological Products | NCT05046548 |

### COVID-19 Trials Tracker

| index | trial_id | registry | registration_date | start_date | retrospective_registration | sponsor |
|---|---|---|---|---|---|---|
| 0 | ChiCTR2000029953 | ChiCTR | 2020-02-17 | 2020-02-01 | True | Zhongnan Hospital of Wul |
| 1 | ChiCTR2000029949 | ChiCTR | 2020-02-16 | 2020-02-16 | False | Zhongnan Hospital of Wul |
| 2 | ChiCTR2000029947 | ChiCTR | 2020-02-16 | 2020-03-01 | False | Longhua Hospital Affiliate University of Traditional C |

### EPPI Centre Covid-19 Living Map of the Evidence

**Tocilizumab in patients admitted to hospital with COVID-19 (RECOVERY): a randomised, controlled, open-label, platform trial**

**Abstract:** <h2>Summary</h2><h3>Background</h3> In this study, we aimed to evaluate the effects of tocilizumab in adult patients admitted to hospital with COVID-19 with both hypoxia and systemic inflammation. <h3>Methods</h3> This randomised, controlled, open-label, platform trial (Randomised Evaluation of COVID-19 Therapy [RECOVERY]), is assessing several possible treatments in patients hospitalised with COVID-19 in the UK. Those trial participants with hypoxia (oxygen saturation <92% on air or requiring oxygen therapy) and evidence of systemic inflammation (C-reactive protein ≥75 mg/L) were eligible for random assignment in a 1:1 ratio to usual standard of care alone versus usual standard of care plus tocilizumab at a dose of 400 mg–800 mg (depending on weight) given intravenously. A second dose could be given 12–24 h later if the patient's condition had not improved. The primary outcome was 28-day mortality, assessed in the intention-to-treat population. The trial is registered with ISRCTN (50189673) and ClinicalTrials.gov (NCT04381936). <h3>Findings</h3> Between April 23, 2020, and Jan 24, 2021, 4116 adults of 21 550 patients enrolled into the RECOVERY trial were included in the assessment of tocilizumab, including 3385 (82%) patients receiving systemic corticosteroids. Overall, 621 (31%) of the 2022 patients allocated tocilizumab and 729 (35%) of the 2094 patients allocated to usual care died within 28 days (rate ratio 0·85; 95% CI 0·76–0·94; p=0·0028). Consistent results were seen in all prespecified subgroups of patients, including those receiving systemic corticosteroids. Patients allocated to tocilizumab were more likely to be discharged from hospital within 28 days (57% <i>vs</i> 50%; rate ratio 1·22; 1·12–1·33; p<0·0001). Among those not receiving invasive mechanical ventilation at baseline, patients allocated tocilizumab were less likely to reach the composite endpoint of invasive mechanical ventilation or death (35% <i>vs</i> 42%; risk ratio 0·84; 95% CI 0·77–0·92; p<0·0001). <h3>Interpretation</h3> In hospitalised COVID-19 patients with hypoxia and systemic inflammation, tocilizumab improved survival and other clinical outcomes. These benefits were seen regardless of the amount of respiratory support and were additional to the benefits of systemic corticosteroids. <h3>Funding</h3> UK Research and Innovation (Medical Research Council) and National Institute of Health Research.

**Author(s)** ;

**Journal** The Lancet

Full record

Coding Report:

Topic

- Treatment evaluation

Version

- All versions

Previous versions 81 to 100

- Version 90 - 30th January 2022

**Evidence Finder**

### COGNITIVE BIAS MODIFICATION FOR SOCIAL ANXIETY: THE DIFFERENTIAL IMPACT OF MODIFYING ATTENTIONAL AND/OR INTERPRETATION BIAS

Yeung, E. S., Sharpe, L.

Cognitive Therapy and Research., 43 : 781 - 791

- Year: 2019
- Problem: Social phobia (social anxiety disorder)
- Type: Randomised controlled trials
- Stage: At risk (indicated or selected prevention)

---

**Evidence Map**

#### Superiority of Step-up Approach vs Open Necrosectomy in Long-term Follow-up of Patients With Necrotizing Pancreatitis

Hollemans et al.
Gastroenterology.
2019;156(4):1016-1026

**Conclusion**

In an analysis of long-term outcomes of trial participants, we found the step-up approach for necrotizing pancreatitis to be superior to open necrosectomy, without increased risk of reinterventions.

**Overall risk of bias judgement**

Some concerns

**Funding**

Non-industry funded

### IDDO

#### The effect of zinc supplementation on inflammatory responses and total blood cells counts of patients who infected with COVID-19

Country
Iran (Islamic Republic of)

Unique trial registry identifier
IRCT20210204050238N1

Date the clinical trial record was first registered with the source clinical trial registry
2021-03-02

Additional unique registration identifiers if the trial was registered with multiple clinical trial registries
-

Short hand reference or acronym assigned to the study. E.g. NOSO-COR; SOLID-C19; SOLIDARITY
-

Source
Iranian Registry of Clinical Trials

Recruitment Status
Completed

### MetaEvidence breast cancer

**I-SPY2, 2020** randomized controlled trial    Risk of bias high

JAMA Oncol 2020 May - Cancer 2014; 120:773 - JAMA Oncol 2020 Sep 1;6(9):1355-1362. - NCT01042379

| Treatments | Patients | Method |
|---|---|---|
| pembrolizumab plus paclitxel followed by doxorubicin plus cyclophosphamide (n=69)<br>200mg intravenous pembrolizumab every 3weeks for 4 cycles plus standard NACT: 80 mg/m2 intravenous paclitaxel weekly for 12 weeks, followed by 4 cycles of 60 mg/m2 doxorubicin plus 600 mg/m2 intravenous cyclophosphamide every 2 to 3 weeks (AC).<br>vs.<br>paclitaxel followed by doxorubicin plus cyclophosphamide (n=181)<br>standard NACT: 80 mg/m2 intravenous paclitaxel weekly for 12 weeks, followed by 4 cycles of 60 mg/m2 doxorubicin plus 600 mg/m2 intravenous cyclophosphamide every 2 to 3 weeks (AC).<br>If patients had an infusion reaction despite corticosteroid premedications, switching to nab-paclitaxel was allowed. | Patients with ERBB2 (formerly HER2)-negative breast cance, women treated with neoadjuvant for early-stage breast cancer (stage II or III)<br>stage II or III breast cancer and primary tumors larger than 2.5 cmby clinical examination or larger than 2.0cmby imaging | Blindness: open label<br>inclusion period: November 26, 2015, and November 5, 2016,<br>Sample size: 69/181<br>Primary endpoint: pCR<br>FU duration: 33.6 vs 42<br>Sponsor: Quantum Leap Healthcare Collaborative<br>P2 /two sided 0,05; BAYESIAN STUDY |

! the study did not meet its primary endpoint pCR

exploratory study (phase 2)

trial #5132 MA 235

### Meta-Evidence COVID

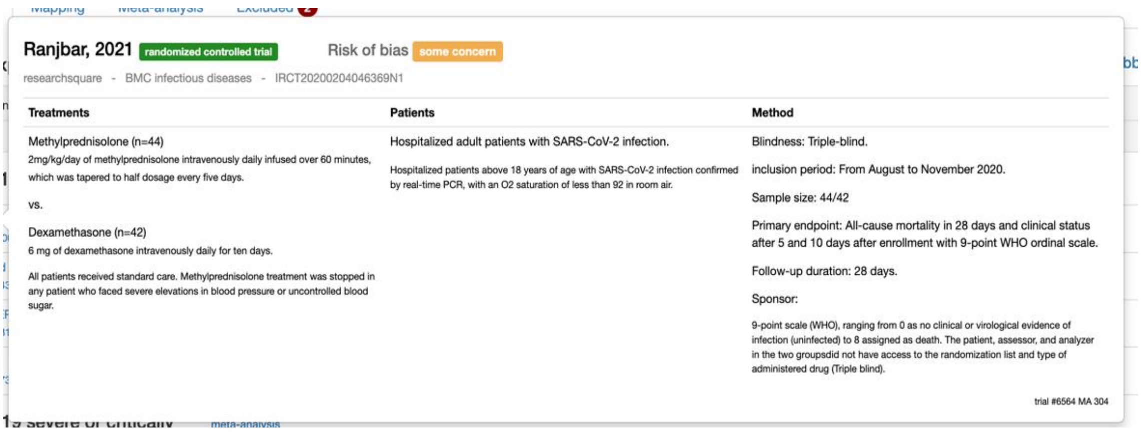
Ranjbar, 2021 randomized controlled trial Risk of bias some concern

researchsquare - BMC infectious diseases - IRCT20200204046369N1

| Treatments | Patients | Method |
|---|---|---|
| Methylprednisolone (n=44)<br>2mg/kg/day of methylprednisolone intravenously daily infused over 60 minutes, which was tapered to half dosage every five days.<br>vs.<br>Dexamethasone (n=42)<br>6 mg of dexamethasone intravenously daily for ten days.<br>All patients received standard care. Methylprednisolone treatment was stopped in any patient who faced severe elevations in blood pressure or uncontrolled blood sugar. | Hospitalized adult patients with SARS-CoV-2 infection.<br>Hospitalized patients above 18 years of age with SARS-CoV-2 infection confirmed by real-time PCR, with an O2 saturation of less than 92 in room air. | Blindness: Triple-blind.<br>inclusion period: From August to November 2020.<br>Sample size: 44/42<br>Primary endpoint: All-cause mortality in 28 days and clinical status after 5 and 10 days after enrollment with 9-point WHO ordinal scale.<br>Follow-up duration: 28 days.<br>Sponsor:<br>9-point scale (WHO), ranging from 0 as no clinical or virological evidence of infection (uninfected) to 8 assigned as death. The patient, assessor, and analyzer in the two groupsdid not have access to the randomization list and type of administered drug (Triple blind). |

trial #6564 MA 304

### MetaIO

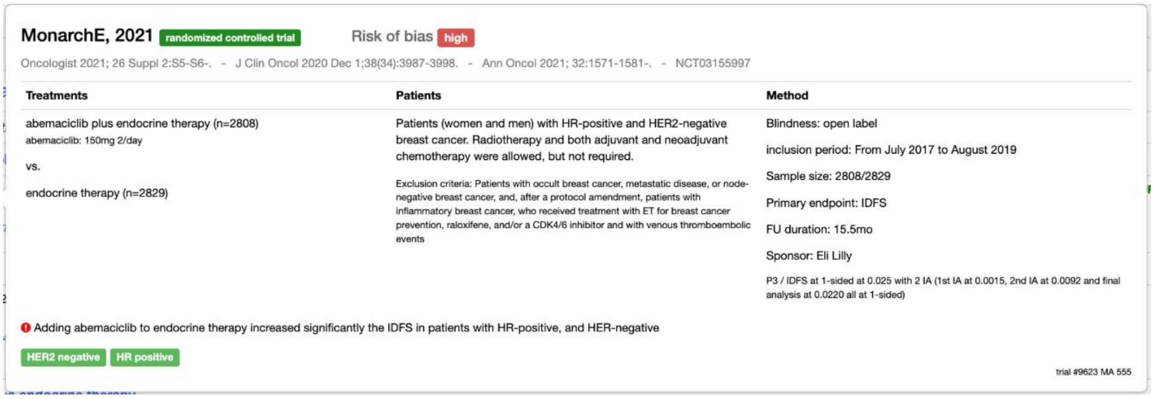
MonarchE, 2021 randomized controlled trial Risk of bias high

Oncologist 2021; 26 Suppl 2:S5-S6-. - J Clin Oncol 2020 Dec 1;38(34):3987-3998. - Ann Oncol 2021; 32:1571-1581-. - NCT03155997

| Treatments | Patients | Method |
|---|---|---|
| abemaciclib plus endocrine therapy (n=2808)<br>abemaciclib: 150mg 2/day<br>vs.<br>endocrine therapy (n=2829) | Patients (women and men) with HR-positive and HER2-negative breast cancer. Radiotherapy and both adjuvant and neoadjuvant chemotherapy were allowed, but not required.<br>Exclusion criteria: Patients with occult breast cancer, metastatic disease, or node-negative breast cancer, and, after a protocol amendment, patients with inflammatory breast cancer, who received treatment with ET for breast cancer prevention, raloxifene, and/or a CDK4/6 inhibitor and with venous thromboembolic events | Blindness: open label<br>inclusion period: From July 2017 to August 2019<br>Sample size: 2808/2829<br>Primary endpoint: IDFS<br>FU duration: 15.5mo<br>Sponsor: Eli Lilly<br>P3 / IDFS at 1-sided at 0.025 with 2 IA (1st IA at 0.0015, 2nd IA at 0.0092 and final analysis at 0.0220 all at 1-sided) |

Adding abemaciclib to endocrine therapy increased significantly the IDFS in patients with HR-positive, and HER-negative

HER2 negative HR positive

trial #9623 MA 555

### MetaPreg

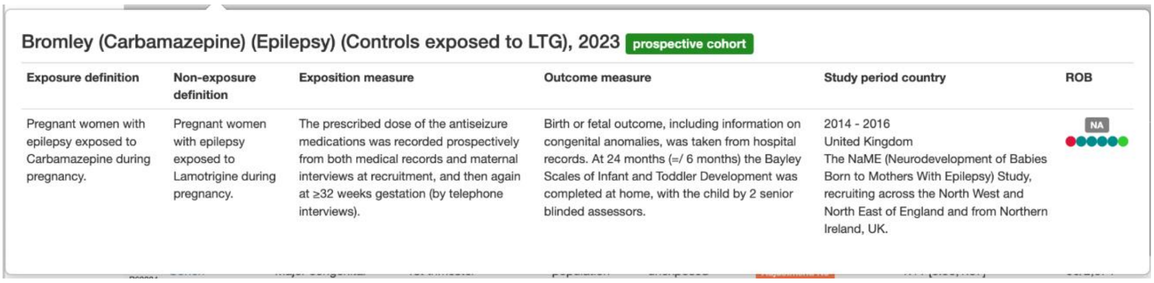
Bromley (Carbamazepine) (Epilepsy) (Controls exposed to LTG), 2023 prospective cohort

| Exposure definition | Non-exposure definition | Exposition measure | Outcome measure | Study period country | ROB |
|---|---|---|---|---|---|
| Pregnant women with epilepsy exposed to Carbamazepine during pregnancy. | Pregnant women with epilepsy exposed to Lamotrigine during pregnancy. | The prescribed dose of the antiseizure medications was recorded prospectively from both medical records and maternal interviews at recruitment, and then again at ≥32 weeks gestation (by telephone interviews). | Birth or fetal outcome, including information on congenital anomalies, was taken from hospital records. At 24 months (=/ 6 months) the Bayley Scales of Infant and Toddler Development was completed at home, with the child by 2 senior blinded assessors. | 2014 - 2016<br>United Kingdom<br>The NaME (Neurodevelopment of Babies Born to Mothers With Epilepsy) Study, recruiting across the North West and North East of England and from Northern Ireland, UK. | NA |

### TrialsResultsCenter

Not working properly.

**WWARN**

| | | B | C | D | E | F | G | H | I | J | K | L | M | N |
|---|---|---|---|---|---|---|---|---|---|---|---|---|---|---|
| 1 | Nu | Repeat Instru | Repeat Insta | Authors of th | Year of Publi | Title of Publi | Journal of Pu | Journal Volu | Issue Numbe | Pages of Arti | Clinical Trials | Region of Stu | Country of St | Language of |
| 2 | 12936-022-04143-4 | | | Pembet Sing | 2022 | Prevalence o | Malaria journ | 21 | 1 | 137 | ACTRN12616 | Africa | Congo (Demo | English |
| 3 | 12 | Treatment | 1 | | | | | | | | | | | |
| 4 | 12 | Treatment | 2 | | | | | | | | | | | |
| 5 | 12 | Site | 1 | | | | | | | | | | | |
| 6 | 12936-022-04350-z | | | Assefa, A.; H | 2022 | Therapeutic | Malaria journ | 21 | 359 | Jan/13 | Not Provided | Africa | Ethiopia (ETH | English |
| 7 | 12 | Treatment | 1 | | | | | | | | | | | |
| 8 | 12 | Treatment | 2 | | | | | | | | | | | |
| 9 | 12 | Treatment | 3 | | | | | | | | | | | |
| 10 | 12 | Treatment | 4 | | | | | | | | | | | |
| 11 | 12 | Site | 1 | | | | | | | | | | | |
| 12 | 12 | Site | 2 | | | | | | | | | | | |
| 13 | 12936-022-04380-7 | | | Hussein, Mol | 2022 | Therapeutic | Malaria journ | 21 | 351 | Jan/13 | Not Provided | Africa | Ethiopia (ETH | English |
| 14 | 12 | Treatment | 1 | | | | | | | | | | | |
